# Understanding the Experiences and Needs of Participants in Decentralised Clinical Trials: A Qualitative Analysis

**DOI:** 10.64898/2026.01.12.26343978

**Authors:** Eoin Gamble, Ciara Heavin, Conor Linehan

**Affiliations:** Applied Psychology, University College Cork, Ireland; Cork University Business School, University College Cork, Ireland

**Keywords:** Decentralised trials, DCT, Remote trials, Patient facing technology, Participant experience, Patient experience, Patient centeredness, Patient Engagement, Participant-centric trials, Human centred

## Abstract

**Background:** Experts across industry, academia, and regulatory bodies increasingly stress the importance of prioritising participant experiences in clinical trials. Simultaneously, the shift towards decentralised approaches is reshaping how trials are designed and conducted, with a direct impact on the experiences of the participants involved.

**Objective:** This study aims to gain a deeper understanding of the experiences of participants taking part in Decentralised Clinical Trials, focusing on the unique challenges and opportunities presented by remote trial participation, and the tools and organisational processes that support participation.

**Methods:** This exploratory qualitative study involved interviews with fourteen individuals who had personal experience participating in remote or decentralised clinical trials. The interview transcripts were analysed using reflexive thematic analysis to identify key patterns and insights.

**Results:** We generated six main themes: (1) Positive experiences with remote participation, (2) co-ownership and meaningful engagement, (3) increasingly fragmented care and support, (4) confusion and uncertainty in navigating the trial, (5) need for human interaction and connection, (6) the hidden workload - navigating the burdens of remote trial participation. While the remote trial is an adaptable and sustainable model offering several benefits for participants and potential for clinical trials, it introduces complexities and challenges for participants that can negatively impact engagement and experience.

**Conclusion:** This study recognises both the potential benefits and challenges introduced by remote trial participation. It underscores the need for technology that not only streamlines trial processes but also responds to the needs, experiences, and well-being of participants. This requires rethinking trial technology features and functionality, strengthening collaboration with stakeholders, and focusing on elements of the participant experience that go beyond remote access and rudimentary data collection.

## Introduction

Clinical trials are essential for testing new treatments and advancing medical understanding(1). They traditionally involve participants making frequent in-person visits to clinical sites(2). Recently, trials have increasingly adopted technologically mediated approaches that reduce site visits by supporting recruitment, intervention delivery, study visits, outcome assessment, and data collection(3–6). The use of such technology is intended to enhance convenience and flexibility for participants(7,8), to increase efficiency, reduce burden(9), and enable passive data collection(10). Terms like decentralised(11), remote(5), direct-to-participant, virtual(6), and hybrid trials that blend decentralised and site-based components(12) have emerged to describe these approaches. For clarity, in this paper, we refer to ‘remote,’ ‘virtual,’ ‘hybrid’ trials, or ‘Decentralised Trials’ collectively as Decentralised Trials (DCTs). Participant retention remains a challenge in remote studies(13). To date, there is limited evidence on patients’ experiences in remote or decentralised clinical trials(14). As these models become more common, understanding how participants experience them is increasingly important.

Recent studies have set out to understand people’s experiences when engaging with DCT approaches. For example, telehealth and remote monitoring are shown to be generally acceptable, with participants valuing reduced clinic visits and the opportunity for daily health monitoring(15). Broader surveys of patients suggest a preference for hybrid models that combine remote and on-site visits but suggest a need to personalise trial organisation to patient needs and conditions(16). Some patients report satisfaction with remote visits and electronic data collection, including overall feasibility and low burden(17,18), but concerns persist. For example a survey on patient perspectives of DCTs found that, although participants appreciated the approach, reduced contact with clinical staff could limit engagement and motivation(19). Participants also encounter technical difficulties with videoconferencing and self-monitoring applications, which require additional support and reminders to use effectively(20,21). Challenges with app usability suggest that digital literacy plays a role in shaping participant experience and may impact participation(22). Participants have expressed a desire for tailoring trial delivery to individual preferences and health conditions, and has been shown to increase willingness to participate(16). Regular communication with study staff is important to maintaining trust and engagement in remote settings(23,24). Positive feedback and regular contact with research staff reinforce the value of ongoing support throughout participation(22). Decentralised approaches may require more support mechanisms than traditional trials; they offer the potential to improve patient safety, treatment efficacy, and communication(25), while also reducing the burden on participants(16).

In parallel with increasing technological mediation in clinical trials, regulatory bodies are placing greater emphasis on patient-centeredness and patient-focused drug development(26), that is, designing trials that better reflect the needs, preferences, and lived experiences of participants(27). Patient-centeredness aims to address patients’ needs across various stages of a trial, including design, enrolment, data collection and outcome reporting(28). In healthcare more broadly, the patient experience has been defined as the sum of all interactions, shaped by an organisation’s culture, that influence patient perceptions across the continuum of care(29), and factors that influence participation and engagement throughout the clinical trial(30). Importantly, research suggests that many of the same factors that shape patient experience in traditional trial settings also apply in remote trials, despite differences in format(23). These include compassion, cultural sensitivity, spiritual support, and personalised attention(31). Positive experiences are also linked to interactions with other patients, building trust with the researchers, and expertise of the research team(32). Other contributions include courteous, knowledgeable study staff, clear communication about study procedures(33), and support throughout participation(34). Regular communication, responsiveness, multiple contact methods, and a genuine sense of care all enhance the patient experience(35). Furthermore, participants can face psychological and physical burdens(36), time demands, frequent clinic visits(37), transportation issues, clinic wait times(38) and frequent changes in staff(39). Negative experiences are tied to participant burden, as well as waiting around and infrequent or impersonal communication(35). In contrast, trial designs that reduce participant burden have been shown to improve engagement(40).

Taken together, these findings suggest that many of the core aspects shaping patient experience in traditional trials remain relevant in decentralised settings. However, the remote nature and greater reliance on technology may alter how these factors are encountered and addressed. This study specifically examines how digital tools and remote approaches affect participants’ needs, motivation, perceptions, sense of support, and relationships with the research team. These factors are central to engagement, retention, and overall trial success(41). While there is growing insight into DCT dynamics, ongoing calls for research(42,43) highlight the need to better understand how decentralised models can more effectively support meaningful, participant-centred participation. Although research has explored participants’ needs in traditional trials, there is limited understanding of their lived experiences in decentralised trials, particularly regarding how technology-mediated interactions and remote engagement shape motivation, support, and relationships with research teams. This study addresses these gaps by drawing on semi-structured interviews with former participants, providing first-hand accounts of how participants experience decentralised trials and how their needs, motivations, and relationships are influenced by remote models. For conceptual clarity, we use the term participants to refer to both trial patients and participants throughout this paper.

## Methods

### Research Design

We conducted a qualitative study using semi-structured interviews to explore participants’ experiences in remote trials that utilised decentralised, technology-driven methods for trial conduct. Our questions focused on the remote nature and how the use of technology in these trials impacted the participant experience. Semi-structured interviews were used to gather data, allowing remote trial participants to share their perspectives(44), and provide first-hand accounts(45) of participants’ experiences. Reflexive Thematic Analysis (RTA), as described by Braun and Clarke(46), was employed to identify meaningful patterns in the data, themes and provide insights into participants’ perspectives.

### Participants

Participants were recruited for their direct experience relevant and informative to the study’s aim of exploring participation in remote, hybrid, and decentralised clinical trial formats(47). To ensure participants met the study’s eligibility criteria, an online screening questionnaire was developed using the Qualtrics platform (see Supplementary Material 3). This questionnaire assessed participants’ eligibility for the study. Individuals were eligible if they were over 18 years of age and had direct experience participating in a remote, hybrid, or decentralised clinical trial. The questionnaire included both multiple-choice and open-ended questions to capture key demographic information and relevant trial experience. Participant recruitment online was supported by individuals within patient advocacy networks, who facilitated introductions to people with relevant experience and provided guidance on online communities suitable for promoting the study. Advocates and participants in our study shared the online screening questionnaire within relevant communities and personal networks, helping to expand the study’s reach. This approach enabled access to individuals with a range of backgrounds, perspectives, and experiences of remote trials. Engaging with advocacy networks ensured that recruitment was informed by those with insight into patient communities, supporting the inclusion of experienced participants who might not typically be reached through conventional recruitment strategies.

Participants were recruited from a range of countries, including Italy, Kenya, the Czech Republic, the United States, England, Switzerland and Scotland, who had been involved in various types of studies, including commercially sponsored and investigator-initiated trials. All participants had prior experience with remote trial formats and had interacted with technologies such as patient apps, electronic diaries, patient monitoring tools, and telehealth solutions. The study adopted a broader, multi-study recruitment approach for two main reasons. First, including participants from different countries and trial types allowed for capturing experiences across diverse systems, cultural contexts, and technological infrastructures. Second, recruitment challenges made it difficult to enrol sufficient participants from any single trial, necessitating a pragmatic strategy that drew on individuals across multiple studies. The resulting sample was intentionally heterogeneous, spanning clinical areas and technologies, which provided a wide range of perspectives. However, this diversity also limited the ability to draw general conclusions, as participants experiences were shaped by different technologies, protocols, durations, and health contexts. Table 1 provides an overview of the study participant group.

**Table 1.** Summary of Study Participants. Overview of interviewed participants (all names are pseudonyms).

| Name | Gender | Age Range | Trial Clinical Area | Completed Trial | Trial Duration | Technology used | Country |
| --- | --- | --- | --- | --- | --- | --- | --- |
| Mary | Female | 54-64 | Oncology | Yes | 3 months | Electronic Diaries / Patient Portals | Italy |
| Celia | Female | 45-54 | Rare Diseases | Yes | 12 months | Electronic Diaries / Phone calls | Czech Rep |
| Jordan | Female | 18-24 | Behavioural intervention trial | Yes | 12 months | Telemedicine / Remote Monitoring Devices | USA |
| Robin | Female | 55-64 | Neurology | Yes | 5 months | Telemedicine / Remote Monitoring Devices / Electronic Diaries | England |
| Mia | Female | 45-54 | Respiratory | Yes | Ongoing | Remote Monitoring Devices / Electronic Diaries | England |
| Cassie | Female | 55-64 | Gastroenterology | Yes | 12 months | Telemedicine / Electronic Diaries | Scotland |
| Mark | Male | 55-64 | Behavioural intervention trial | Yes | 12 months | Telemedicine / Patient Portal / Patient app | England |
| Jessica | Female | 65+ | Respiratory | Yes | 3 months | Remote Monitoring Devices / Electronic Diaries | England |
| Emma | Female | 55-64 | Respiratory | Yes | Full Trial Duration | Telemedicine / Electronic Diaries / Patient app | England |
| Rose | Female | 35-44 | Dermatology | Yes | 6 Months | Telemedicine / Patient Portal / Patient app | Kenya |
| Helen | Female | 35-44 | Endocrinology | Yes | 2 months | Telemedicine / Electronic Diaries | Switzerland |
| Lilly | Female | 45-54 | Immunology | Yes | 12 months | Electronic Diaries / Patient Portal | Switzerland |
| Kim | Female | 65+ | Respiratory | Yes | 6 months | Remote Monitoring Devices | England |
| Sonya | Female | 45-54 | Behavioural intervention trial | Yes | 12 months | Remote Monitoring Devices / Electronic Diaries | England |

### Data collection

Data collection took place between September 2024 and March 2025. To accommodate participants’ schedules and locations, interviews were held online via Microsoft Teams video calls. Each interview, conducted by the first author, lasted approximately one hour. The interview guide was designed based on a review of the literature to ensure alignment with the research question (see Supplementary Material 1). The questions were drawn from existing literature(29) and focused on understanding participants’ experiences with the remote nature of the trial, their interaction with technology, the trial’s conduct, the participant-clinician relationship, the support received, and their specific needs. A collaborative process took place between the research team to refine and enhance the interview guide. A conversational approach was used during the interviews, incorporating open-ended(48) and follow-up probe questions(49), allowing interviewees to elaborate on their experiences. The interviews were recorded and transcribed via Microsoft Teams, with the first author verifying their accuracy. After completing fourteen interviews, and in line with Braun and Clarke’s subjective view that data saturation is a context-dependent, interpretative judgment(50), it was determined that sufficient data had been collected to effectively address the research question.

### Data Analysis

The authors followed the six-phase Reflexive Thematic Analysis (RTA) framework outlined by Braun and Clarke(46). The first author was responsible for independently coding the data, identifying relevant codes and themes that aligned with the research objectives. A collaborative refinement process took place through discussions with co-authors, ultimately reaching an agreement on the final interpretations. In Phase 1, the first author became familiar with the data by transcribing and listening to the recorded interviews and reading and re-reading the transcripts and documenting initial thoughts and observations. Phase 2 entailed generating initial codes through a systematic analysis of the data, to detect significant patterns, a time-consuming process that required reflection to keep them precise and aligned with the research question. In Phase 3, codes were grouped into potential themes and refined through an iterative process, revisiting codes and transcripts to ensure they stayed clear, specific, and closely tied to the participants’ data. Phase 4 involved reviewing and refining themes to ensure alignment with the data, with collaborative discussions helping to challenge interpretations and define coherent overarching themes and sub-themes. Phase 5 involved final refinement, where themes were clearly defined, named, and organised into main themes and sub-themes through collaborative discussions, ensuring titles accurately reflected participants’ experiences. In Phase 6, the analysis was presented, with each theme supported by detailed descriptions and participant quotes, ensuring that the findings were rooted in the participants’ experiences. Examples of the data analysis are provided in Supplementary Material 2

### Ethical Considerations

The University Social Research Ethics Committee (SREC) granted approval for the study in June 2024. Participation in the study was voluntary, and participants received electronic information sheets and consent forms that clearly explained the study’s purpose and procedures during the recruitment process. Personal details were anonymised, sensitive information was removed from transcripts, and participants were informed of their right to withdraw at any stage. No compensation was provided, ensuring participation was motivated solely by interest in contributing to the research.

### Findings

Our findings are structured around six themes: (1) Positive experiences with remote participation, (2) Co-ownership and meaningful engagement, (3) Increasingly fragmented care and support, (4) confusion and uncertainty in navigating the trial, (5) Need for human interaction and connection, (6) and the hidden workload - navigating the burdens of remote trial participation. Each theme has several sub-themes, as illustrated in Figure 1.

**Fig 1.**
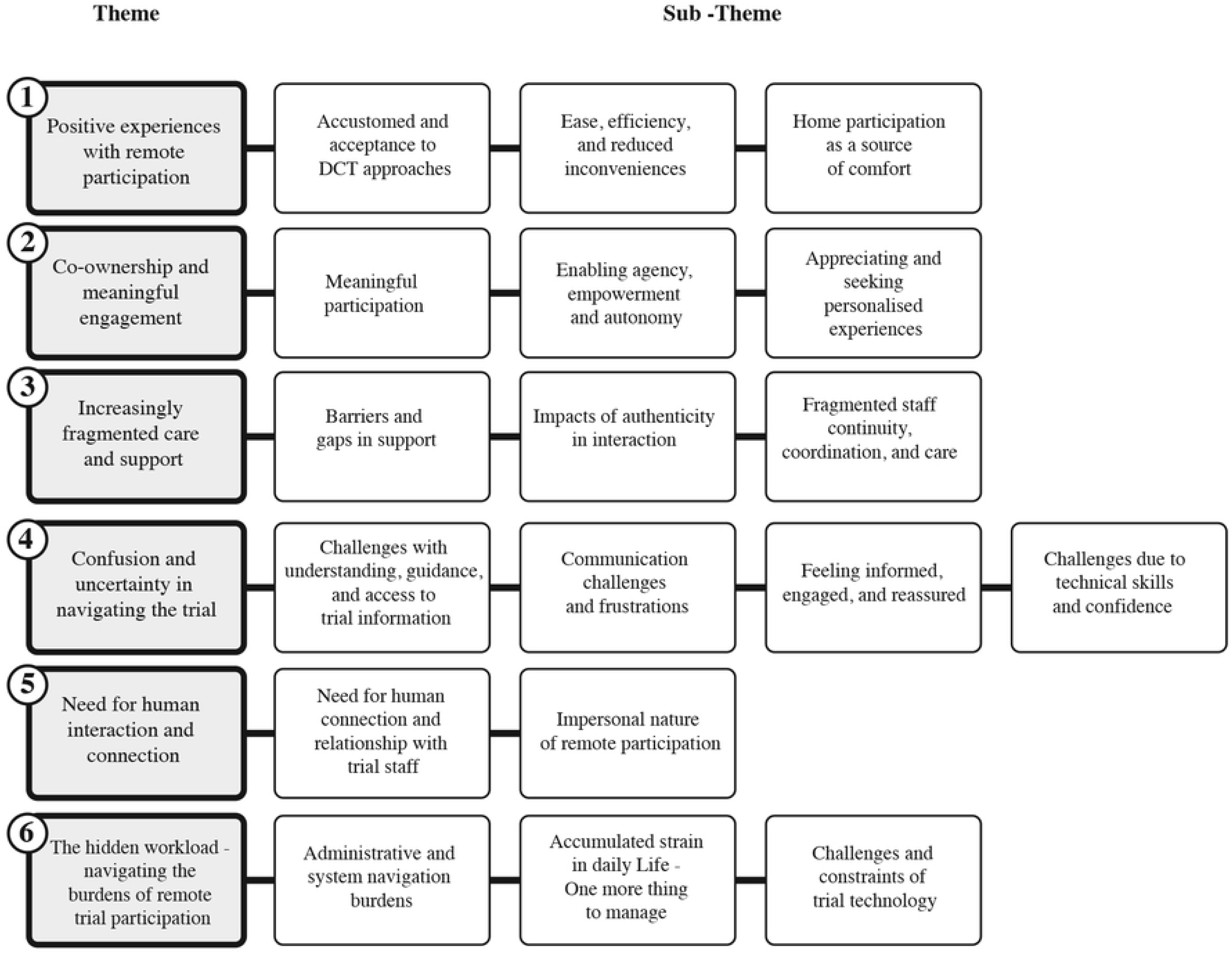
Findings – Themes and Sub-themes. Illustrates the key themes and sub-themes that developed from the study.

## Theme 1 - Positive experiences with remote participation

This theme captures participants’ generally positive experiences with elements of remote trials, particularly the benefits of easier access and fewer barriers to participation. Completing trial activities from home was seen as convenient, helping them avoid the logistical and physical challenges of in-person visits. Overall, participants were familiar with remote formats and viewed them as an acceptable and practical way to engage with a trial.

### Sub theme - Accustomed and acceptance to DCT approaches

There was a broad acceptance and in some cases a preference for remote approaches in trials. This was influenced by several factors. Some participants’ were familiar with remote working and technology use, particularly through their jobs. To some, remote monitoring and trial tasks were seen as routine and manageable. Some viewed remote monitoring as a suitable approach for managing chronic conditions. Other participants, particularly those less familiar with technology, described initial hesitation, but they became more comfortable over time. Overall, participants supported and some even advocated for the continued use of remote methods in clinical research.

> *“So actually, what we did on the trial was totally normal for me. It’s how I’ve operated for the last eight years.”***- Mark**

> *“I didn’t realise there’d be so many apps to go with it. But it’s actually fine. Once you understand what each one’s for, and once you’ve done it a couple of times, it’s so easy.”* **- Mia**

### Sub theme - Ease, efficiency, and reduced inconveniences

Participants described the remote trial as manageable and accessible, highlighting the removal of inconveniences like travel, public transport, and needing to navigate hospital environments. This not only saved time and lowered personal costs but also eased the physical and health-related strain of attending a site. Compared to paper-based methods, participants discussed how remote approaches reduced both cognitive and administrative burdens, requiring less effort. Patient-facing technology played a key role by simplifying involvement, supporting adherence through convenient data collection, and enabling efficient participation without disruption. Flexible scheduling options further contributed to a smoother experience, allowing participants to reschedule easily when needed. Additionally, participants appreciated access to upgraded medical equipment for use at home, enhancing their overall engagement.

> *“I didn’t need to leave the house to go to a study site. I didn’t have to brave public transport. I didn’t have to brave car parking. Sick people in the hospital, any of that. It didn’t need me to go to hospital. It takes two hours each way, and then there’s an awful lot of sitting around and I work full time. So to go to a study site involves time, expense, and health concerns.”* **- Lilly**

> *“I think the remote nature was brilliant because it meant that you weren’t having to traipse up to London several times to go and blow into bits of kit once or twice to see what happens.”* **- Kim**

### Sub theme - Home participation as a source of comfort

Home-based participation was linked to feelings of flexibility, privacy, low pressure, and enhancing the overall experience. Beyond convenience, it offered comfort and a relaxed setting that helped normalise the process and put participants at ease. However, one participant pointed out that the relaxed home environment could sometimes reduce motivation.

> *“I’m at my own home, dressed the way that I want. Don’t have to get prepared to go out into public…. it’s more flexible because I’m at my own home and I’m in a place that I feel comfortable and so it generally feels positive to me… I’m safe here. I can talk to the researchers, and I can do it in my private space.”* **- Jordan**

### Theme 1 Discussion

Similar to Davies and Slykerman(51), our research highlighted how participants generally accepted decentralised methods. However, we also found that their initial level of comfort varied depending on their technological competency. Those with more experience, such as remote workers, adapted and valued the technology, while others initially struggled, but comfort generally improved over time. These findings are consistent with previous research indicating that varying levels of digital familiarity among stakeholders can influence the ease of participation in decentralised clinical trials(7). Notably, participants viewed DCT model positively, as it helped lower barriers to participation. Past research has highlighted several barriers in traditional trials, including time commitments, frequent clinic visits(37), transportation issues, and long waiting times at sites(38), all of which contributed to participant frustration(35). Logistical hurdles, such as travel time and distance to trial sites, also pose significant challenges to participation(52). Many of these burdens are largely removed in remote trials and appreciated by the participants. Remote activities can fit into daily life. Participants valued the flexibility, better home equipment provided, and reduced administrative burdens compared to paper methods, supporting the idea that DCTs can lower elements of participation burden and support flexibility(3,8). Reduced burden is particularly important for retaining participants in clinical trials(53) and improving engagement(40).

## Theme 2 - Co-ownership and meaningful engagement

This theme highlights the role of participants as active partners in remote clinical trials, emphasising their sense of co-ownership and meaningful engagement. Participants attributed personal significance to their involvement, shaping their motivation and commitment. The trial experience was strengthened when participants felt empowered, with greater agency and autonomy during the process. Additionally, a desire for personalised experiences that align with individual needs and preferences.

### Sub theme - Meaningful participation

Participants described a sense of ownership in the trial, feeling like partners rather than just subjects. While enthusiasm was high at the start, some experienced the relationship as one-sided and sought greater recognition and validation. When their role, knowledge, and contributions were acknowledged, it helped build trust and fostered a stronger connection to the trial.

> *“I tend to just do, I don’t tend to think about myself. I think I manage my situation by being focused on doing. I’m finding meaning in what I’m doing. That’s what works for me.”* **- Sonya**

For some, involvement was also seen as personally meaningful and intellectually rewarding. Participants appreciated the chance to gain new knowledge, insight into their health, and contribute to something larger than themselves. Motivations ranged from curiosity and self-data analysis to altruism, with participants valuing the opportunity to make a societal impact, even when it involved personal sacrifice.

> *“But perhaps 5-10 years down the line, there will be something that will come from that or, either directly from that, or someone, somewhere will then read that and it will set them off in a different direction that will benefit mankind.”* **- Cassie**

For some, the trial technology fostered a sense of inclusion and a connection to the trial. Others found emotional or practical benefits, especially during periods of ill health or social isolation. Some participants saw it as a way to preserve their independence; others appreciated the break from routine that the trial provided. Some found comfort in self-monitoring, the novelty of the technology, or simply the distraction it offered from their health concerns.

> *“It gives me something that I feel like I’m doing something useful, and it gives me something to take my mind off everything else, really. So if I’m doing that twice a day and making sure I’ve done it all properly and whatever, I think that’s good for me.”* **- Mia**

### Sub theme - Enabling agency, empowerment and autonomy

Participants experienced a sense of empowerment through active participation, with autonomy and flexibility enhancing their involvement. This empowerment was strengthened by reviewing feedback and metrics, as well as by familiarity and competence gained through prior experiences, which fostered greater control and agency throughout the trial. Technology played a key role by supporting self-management, allowing participants to work at their own pace and control task completion and decision-making. Participants preferred autonomy in scheduling and access to information, but frustration arose when they felt a lack of control over the technology, highlighting the need to balance flexibility with effective support mechanisms.

> *“I like the autonomy. That was great…. I like the fact that it relied mostly on me.”* **- Sonya**

### Sub theme - Appreciating and seeking personalised experiences

Participants described frustration and disengagement caused by standardised messages, templated approaches, and rigid procedures. Some felt the experience was impersonal and unresponsive to their needs. Where personalisation was lacking, participants sometimes felt emotionally distant or disconnected from the trial.

> *“It was very impersonal. And it’s like bulk mailing from a study team.”* **- Lilly**

> *“And as I said, it was far less personal. It didn’t feel like the people really got to know me at all. It was just perfunctory questionnaires.”* **- Robin**

In contrast, participants highlighted the positivity of a personalised trial experience, especially in communication styles, feedback mechanisms, and flexible protocols. When these elements were tailored to individual needs, participants felt more supported and engaged, reporting greater satisfaction when interactions felt individualised and responsive.

> *“They really made sure that everything was personalised according to my level of knowledge, according to my interest, according to what I already knew. And so that I didn’t have to sit through yet another hour of, for example, education on the material itself. If I already know it, then that is wasted time for both them and me. So it was actually quite smart that way.”* **- Helen**

Personalised care, adaptable technology, and attentive clinical support fostered a sense of being valued, encouraging openness and reinforcing engagement with the trial. Some participants deepened their connection by relating feedback from technology to their physical experiences, which strengthened their sense of ownership and meaning.

## Theme 2 - Discussion

Our research showed that participants’ motivation reflected a layered mix of altruism, personal growth, and both symbolic and material benefits. Participants described a sense of ownership over their experience, viewing themselves as active contributors motivated by validation, impact, and meaningful contribution. This aligns with past research showing that personal meaning and altruism are powerful motivators in clinical trial participation(54), and that intrinsic motivation, rooted in personal values and goals, supports sustained motivation(55), a finding that held true even in the remote context of participants’ trials. Autonomy played a central role in shaping experience. Expressed through flexibility and convenience, it enabled participants to exercise control over how and when they engaged with the trial. But autonomy also introduced new responsibilities that, if unsupported, could become burdensome and undermine motivation. As both an ethical requirement(56) and a psychological need(57), autonomy must be carefully supported through systems that offer clarity, feedback, and reassurance. Personalisation was equally important and aligns with previous research showing that particpants value trials tailored to their individual needs(16). It’s also a key factor shaping trial experiences(31). Participants valued interactions and feedback that acknowledged their individual roles and progress. These personalised touchpoints helped reinforce their sense of agency, purpose, and connection to the trial’s broader goals, factors known to support engagement and retention(41). Yet, in decentralised settings, opportunities for this kind of tailored engagement can be limited(58). In contrast, treating participants as partners improves engagement and better outcomes(59,60).

## Theme 3 - Increasingly fragmented care and support

Participants discussed the fragmented nature of care and support in remote clinical trials. Gaps in support structures and limited clinician understanding of participants’ experiences, disjointed continuity of trial staff, a lack of medical coordination and integrated care further disrupted the trial experience.

### Sub theme - Barriers and gaps in support

Participants reported gaps in support during remote trial participation, including delays in responses, slow resolution of technical issues, and limited, timely, emotional, or clinical input. The lack of immediate assistance led to feelings of uncertainty, isolation, and frustration, with support described as insufficient and reactive. Limited access to clinician guidance and expert advice compounded concerns about responsiveness. Technological platforms were perceived as detached, relying on automated replies and asynchronous email communication, while structural barriers such as restricted communication windows and geographical limitations further hindered real-time problem-solving and access to care. In contrast, when participants had direct access to troubleshooting, emotional support, and post-trial assistance, their experience improved markedly. Responsive technology and self-tracking tools fostered feelings of safety and reassurance, with many emphasising that timely support was a critical component of care in remote trials.

> *“The frustration of not being able to get the support you want when you need it and not being able to see the person when you need them to ask one or two questions when something has come up, you need to talk to someone as soon as possible……. And also just the fact that I have to get someone to help me, and it has to be through an email. Then that feels, oh my God, they’re so far away. You just realise that there’s a big barrier.”* **- Rose**

### Sub theme - Impacts of authenticity in interaction

Participants described how clinical research staff sometimes lacked understanding of their lived experiences, exacerbated by limited feedback channels and the absence of direct clinical observation. Some felt unheard and unable to fully express themselves due to restricted communication methods, limited response options, and reporting constraints. These barriers, amplified by the remote trial setting, increased frustration and created a sense of being misunderstood, which ultimately reduced participants’ confidence in their care. In contrast, when clinicians showed empathy and understanding to participants, it enhanced the overall experience. Opportunities to provide feedback during in-person visits and to be genuinely listened to helped participants feel valued and cared for, highlighting the importance of clinician empathy in the trial process.

> *“It was frustrating because I felt like there was a resource they wanted and they did not really want to hear what the patients say beyond what they wanted to hear.”* **- Rose**

> *I don’t think you were asked to reflect and give feedback regularly enough. And so it was very much driven by the researchers. They’ve got this set idea of what they were going to do and what needed to happen, but they didn’t sort of then get feedback and check that it was working for the people as it went along.” **-*** **Robin**

### Sub theme - Fragmented staff continuity, coordination, and care

Fragmented continuity in trial staffing negatively affected the patient experience, as participants encountered different research staff, leading to a lack of consistency and disjointed communication. This caused frustration.

> *“Because every time there are different people who reply to you, you have to start from the beginning. I thought they would have some kind of file with all my information, but every time it seems like they are reading this information for the first time.”* **- Mary**

Poor coordination of care further impacted participants, who faced fragmented medical support and had to navigate their care independently. Limited interaction between research staff and family doctors, along with delegation of care to external providers, made it difficult for participants to connect with the relevant medical team, resulting in a sense of detachment and increased burden. In some cases, the absence of timely expert intervention, especially during emergencies, left participants feeling unsupported and added to their stress.

> *“Then I would have to fill out the questionnaire that things are not going well, and sometimes I had to seek medical help pretty quickly, and that was more stressful and difficult to arrange…… they would just call me and say you should go to the doctor and ask him to do these blood tests or this analysis. If you’re feeling bad.”* **- Celia**

In contrast, when continuity and coordination were present, participants felt better supported, which enhanced their overall trial experience.

## Theme 3 - Discussion

Our findings suggest that DCTs can reduce research staff oversight and limit their ability to respond effectively to participant needs. This reduction in support contributed to frustration among participants, who sometimes experienced a sense of inadequate care. These results align with Gamble et al.(58), who describe similar challenges in supporting participant needs across decentralised settings. Participants in our study also reported that fragmented care, disconnected support systems, and limited coordination created both emotional and practical burdens, leaving them to manage issues alone. These gaps in care served as a stressor and impacted their overall experience. The importance of a sense of care(35) and support(34) are critical to supporting the patient experience. Integrated and continuous care is often highlighted as an important aspect of patient-centred care(61) and effective patient care requires responsiveness to individual needs(62,63). However, staff changes(39) can disrupt key elements such as continuity, support, and medical coordination, all of which are especially important in decentralised clinical trials(64,65). The remote nature of DCTs intensifies these challenges by limiting in-person availability and immediate support. While remote models may offer flexibility and autonomy, without accessible, empathetic systems, participants can experience these responsibilities as burdensome, resulting in feelings of disconnection, abandonment, and emotional strain.

### Theme 4 - Confusion and uncertainty in navigating the trial

This theme highlights the challenges participants faced in understanding various aspects of the trial. Ambiguity, a lack of clear guidance, and limited access to educational resources contributed to confusion, uncertainty, and difficulty navigating the trial process. Communication challenges further compounded issues, and participants sought but did not always receive feedback and reassurance. Additionally, varying levels of technical confidence presented further obstacles.

### Sub theme - Challenges with understanding, guidance, and access to trial information

Some participants described their experience in remote trials as a “black box,” reflecting a lack of clarity and understanding about how the trial worked, their role, and the part technology played in data collection. This ambiguity limited their engagement, increased frustration, and sometimes led them to question their own understanding and abilities, causing stress and uncertainty. These issues could be heightened in remote settings, when participants struggled to grasp trial processes and expressed a need for more structured guidance.

> *“I didn’t quite see how they would be able to correlate my data on a daily basis to how I was feeling and then what I was thinking, and you know all of these things. honestly, I didn’t see the point of it and how it would relate to the other data.”* **- Helen**

This lack of clarity was compounded by difficulties in accessing and understanding trial information. Some participants struggled with the clarity of the materials provided and digital barriers that hindered comprehension. Difficulties in navigating online information further compounded these issues, limiting their ability to engage effectively with the trial process and increasing stress.

> *“All the information about this medication is online. But sometimes that information online it tries to be very bullet points and then sometimes it’s not easy to understand.”* **- Mary**

Participants highlighted a need for more structured training and clear user instructions. Inadequate induction and broad guidelines left many feeling unprepared and unsupported, undermining confidence and engagement. Foundational knowledge was seen as crucial to reducing uncertainty and lowering barriers to participation.

> *“I think that’s the poor induction, if I’m honest. I think also, we did a follow up call after about a week. and that was when she said by the way, all the other people I have spoken to did not realise that you have to fill in this.”* **- Jessica**

> *“But it was very limited training because at the beginning it seems easy… I thought that was enough, but then in the real moments was not enough.” **-*** **Mary**

### Sub theme - Communication challenges and frustrations

Participants faced barriers in reaching the right person and needed to make extra effort to connect with appropriate staff. Limited communication channels, combined with the absence of nonverbal cues in remote settings, made engagement more difficult and left some feeling frustrated. Ineffective communication, such as delays, gaps in responses, and automated replies, led to a sense that their concerns weren’t being fully addressed, which in turn contributed to disengagement and diminished the overall patient experience. This lack of responsiveness also reinforced a preference for in-person interactions, which were seen as clearer and more supportive.

> *“I had to take a list of number of phones and try to phone everyone to try to find a solution ….. And the first no reply, second no reply, this was difficult for me…. In the moment that you need to quickly reply, this is a problem It seems like when you call, it’s to a call centre. There is something automatic and they reply to you in an automatic way, without really understanding the situation.”* **- Mary**

### Sub theme - Feeling informed, engaged, and reassured

Some participants described how technology-enabled feedback and visual metrics supported their engagement, boosted confidence, and made the experience more enjoyable. For some, the technology provided feedback offered insight into their trial journey, creating a sense of progress, involvement, and even perceived health improvements. Participants stressed the importance of feedback being dynamic, meaningful, and personalised. Some felt the trial app could better deliver this information, helping to maintain motivation, offer reassurance, and prevent feelings of disconnection. When feedback was delayed or missing, especially at key moments, it left some feeling overlooked, unsupported, and disconnected. A lack of meaningful feedback and information, both during and after the trial, led some to feel unimportant or forgotten, particularly when they weren’t updated on their contributions or the outcomes of the research.

> *“If I could see the data points, the data collection points, and what’s to come, that would be great in the study, providing much more guidance. … I would have loved to have known that my blood sample made it each time, not the results, because I knew it took time to process. But I would have loved to have known that they’d got what I’d sent them.”* **- Lilly**

### Sub theme - Challenges due to technical skills and confidence

Participants experienced challenges related to varying levels of technical skills and confidence. Some felt intimidated by the technology provided, struggling with setup or specific devices, and some would have preferred paper-based instructions. There was a contrast between initial impressions of patient-facing technology and the reality of using it in practice, which created further challenges. Although the systems seemed easy at first, difficulties arose once the trial began. Some participants struggled using the technology as expected, which affected their ability to meet trial expectations and led to feelings of intimidation and low confidence.

> *“I suppose initially, when we were trying to set up the technology, I did think, oh no, this is a bit intimidating. If I was in the room with this person, they could actually just put the information on my iPad, or my phone, and I wouldn’t have to do anything.”* **- Robin**

## Theme 4 - Discussion

We identified challenges in participants’ understanding and training related to the trial, including ambiguity, inconsistent communication, and a lack of structured support. These left some participants feeling unprepared and anxious. Past research on patient retention in clinical trials has identified patient competence as a key factor in engagement and retention(41), competence acts as a core psychological need(55), that drives motivation and ongoing participation in clinical trials. Our findings show how easily this sense of competence can be disrupted and how digital access alone is not enough to support this sense of competence. A clear understanding of study procedures(33) is crucial to reducing uncertainties and enhancing participants’ confidence throughout the trial. Feedback, both data updates and relational, are essential for engagement(41), when delivered empathetically, contextually, and timely it enhances the experience, competence, and strengthening participants’ connection to the trial. Like findings in other DCT research(58), challenges in communication between the research team and participants extended beyond technical support. Clear, timely communication and feedback are central to building participants’ sense of competence. They need reassurance about their role, progress, and contributions through feedback that is empathetic, contextual, and relational. Communication and feedback are not just operational tasks, they are relational tools that convey care, value, and inclusion(35). When responsibility falls entirely on the participant without sufficient understanding or feedback, it can create emotional and practical strain, potentially affecting retention, adherence, and data quality. These challenges are compounded by potential issues of digital exclusion in terms of both people and the technology(66). Assumptions about technological literacy and autonomy can obscure the reality that patients’ skills, confidence, and access vary. Competence, then, is shaped not only by what participants are expected to do, but by how they are supported cognitively, practically, and emotionally.

## Theme 5 - Need for human interaction and connection

This theme highlights an erosion of social interaction in remote trials and participants’ need for meaningful connection. Some participants perceived their involvement as transactional, lacking personal engagement and depth in interactions with trial staff. A sense of disconnect and limited relationship-building diminished their experience, contributing to feelings of isolation. Reduced interaction with trial personnel reinforced this sense of detachment. Participants sought greater human connection and valued opportunities for more personal engagement throughout the trial.

### Sub theme - Need for human connection and relationship with trial staff

Participants’ experiences revealed a lack of meaningful personal connection with trial staff, reflecting a broader sense of disconnection in remote clinical trial settings. Some participants reported not getting to know the research team and a lack of opportunities to build relationships, which contributed to feelings of detachment and a sense of impersonality.

> *“There’s really no relationship. There’s no touch. There is really no assessment, like it’s not really human. You feel like there’s a disconnect.”***- Rose**

Participants expressed a desire for human connection, highlighting how personal interactions contribute to a more supportive and humanised trial experience. They valued in-person contact with research staff, which helped build trust, comfort, and a sense of being respected. Informal conversations and the presence of friendly, familiar staff, especially nurses, offered emotional reassurance and helped participants navigate concerns or uncertainties during the trial. These friendly, informal exchanges fostered trust, reassurance, and a genuine feeling of being cared for.

> *“Even though it is remote and it’s about technology and it’s about data, it’s really the people who I got the chance to interact with and that I got to work with. Like the study nurse that I mentioned before. For example, she was lovely and she was very understanding.”* **- Helen**

Despite being provided with digital tools, participants still relied on human coordination and contact to feel confident and cared for. Participants viewed in-person visits as more thorough and attentive, providing better oversight and emotional support. This helped manage anxiety and strengthened motivation to remain engaged. Participants expressed a desire for more in-person interaction during these trials and suggested participant community-building mechanisms to foster connections during trials.

> *“My experience was that face-to-face I would get much more information. The doctors would get to see me physically as well, you know, a physical examination and so on. So that was the component that during the virtual communication, was missing.”* **- Celia**

### Sub theme - Impersonal nature of remote participation

Participants expressed a desire to be recognised as individuals, but the structure and delivery of remote trials at times left them feeling like data providers rather than people. The experience was described as clinical or inhuman, shaped by impersonal, standardised digital platforms. Digital responses came across as detached, and the remote format was seen as an incomplete substitute for direct human support.

> *“You are more like a robot because you’re communicating with something you don’t really have a relationship with, and you’re just giving - This is happening, This is happening, This is happening. You know, (gesture regarding giving data, giving data, giving data) It’s only like it’s not really human.”* **- Rose**

The lack of face-to-face contact, non-verbal cues, and informal conversation reduced the depth of interaction with trial staff, making appointments feel cold and distant. Some participants felt these interactions were less meaningful than in-person visits. Inconsistent communication and time-zone challenges contributed to a sense of detachment, feeling overlooked or dismissed, and feelings of efficiency prioritised over empathy. These experiences deepened participants’ desire for more human connection and recognition during trial participation.

> *“It did just feel like you entered into taking part in something where you were really revealing personal information and how you felt in your mood, and in some ways, I didn’t find it helpful that it was remote at all. It didn’t work for me. I think I needed to sit with someone in a room and have ease of contact.”* **- Robin**

## Theme 5 - Discussion

Our study found that remote trials can weaken the relational dynamic between particpants and research teams, limiting both personalisation and opportunities for interpersonal connection. Negative experiences centred on impersonal communication(35), with participants in our study describing experiences that felt impersonal or transactional, which could diminish motivation and long-term commitment. However, it is important to recognise that not all remote trials resulted in a lack of connection. In some cases, participants appreciated the opportunities where interpersonal interaction with research staff was part of the process and highlighted the positive impacts of this. Human connection is recognised as central to therapeutic engagement and the relationship between trial staff and participants, playing a vital role in promoting positive experiences in clinical trials(67). Relationships form a critical foundation for participant retention(41), supporting treatment adherence throughout the trial process(68), and fulfil a psychological need for motivation(55). While remote models offer clear logistical benefits, they also risk compromising a core element of patient-centred or person-centred care(69).

## Theme 6 - The hidden workload - navigating the burdens of remote trial participation

This theme highlights the hidden workload for participants associated with remote trial participation, emphasising the burdens placed on participants. Navigating administrative tasks and trial systems added complexity, often requiring significant effort to manage. Challenges with trial technology further compounded these difficulties, creating additional barriers to engagement. Participants felt the responsibilities placed on them and relied on informal or external support networks to cope with these demands. Furthermore, they faced several personal challenges, and trial responsibilities were one more thing for them to manage while also managing their health.

### Sub theme - Administrative and system navigation burdens

Some participants described the administrative burden and complexity of navigating remote trial systems as a source of frustration. Tasks like scheduling, managing communications, and handling study responsibilities felt disjointed and time-consuming. These difficulties were made worse by rigid protocols and inflexible systems that didn’t accommodate participants’ needs or offer clear pathways for assistance. Furthermore, the inconvenience of required travel for in-person visits and tests also impacted the participant experience.

> *“It is frustrating, and that does actually need much more effort from me. Whereas if there was a button to say or question, have you been vaccinated, or are you preparing to be vaccinated, yes or no. And then will you be pausing your medication? Yes, from when? When are you starting the pause, and then when are you picking up again? That would be super helpful.”* **- Lilly**

At times, participants felt that too much responsibility was placed on them, managing tasks, making decisions, and handling emergencies in the absence of sufficient support. Participants expected the systems to be straightforward but found themselves navigating confusing processes alone, which created stress, uncertainty, and a sense of isolation. The combined effect of inflexible procedures and lack of adaptability negatively shaped their experience.

> *“It seems that I have to take the responsibility myself. And this is the thing that makes me more isolated. OK, now I have to make some decisions that maybe I am not an expert to take.”* **- Mary**

### Sub theme - Accumulated strain in daily Life - One more thing to manage

Beyond the challenges of navigating trial systems, participants described the ongoing emotional and practical toll of trial participation as it unfolded alongside daily responsibilities, health fluctuations, and the heightened anxiety and stress caused by illness, as well as other life events. This wasn’t just about any one task being difficult; rather, it was about the gradual build-up of demands over time, which could erode motivation and make trial tasks feel like yet another obligation in an already unpredictable situation. For some, they underestimate the long-term demands and what started as manageable tasks, like completing digital diaries or managing technology, became harder to prioritise due to disease management or the fatigue of long-term involvement. Adherence was sometimes impacted not by intention, but by forgetfulness, stress, or the difficulty of managing multiple demands at once. Trial participation, while important, became “one more thing to manage” in an already complex context of illness and everyday life.

> *“Days when the Crohn’s was particularly bad. And I just didn’t want to move. Or if I thought, I’ve got to be online now, and I might have to run to the toilet. I never did. But that was always in the back of your mind.”* **- Cassie**

### Sub theme - Challenges and constraints of trial technology

Participants described a range of technology-related challenges that negatively affected their experience. Frustration arose from unreliable technology, system glitches, poor connectivity, and usability issues that disrupted task completion and data submission. Some noted that individuals with more serious health conditions might struggle with the technology provided. Expectations for seamless, intuitive digital experiences were at times unmet; some participants described the tools as rigid, outdated, uninspiring, and frustrating to use. Technology issues increased anxiety, impacted mood and motivation, and added to the emotional burden of remote participation. Additional barriers included conflicts between device preferences, platform limitations, and the inconvenience of managing multiple devices. For some, the cumulative demands, frequent data entry, excessive information, and manual assessments led to technological fatigue and reduced engagement.

> *“I was saying I can’t click on this. I would send an email saying I’ve tried this and I can’t get this into the text box, and she would then talk to the technical support, and they would come back and say, have you got the font size increased on the phone? Or will you need to take that off and then do this, and you’re like, oh, what a hassle.”* **- Cassie**

> *“I think at the start, I thought it was easy. It was just like pa, pa, pa, and I’m done. As you move on, it gets overwhelming. It gets a bit tiring. It gets a bit boring. When you started, you think it’s easy. But as you go on, it’s not that easy.”* **- Rose**

There was a reliance on participants’ personal formal, informal, and external support networks in using technology. Some participants deferred decision-making or understanding to family members or other external sources of support. This reliance created a passive role for participants, highlighting the reliance on family navigating the complexities of trial technologies.

> *“I had my youngest with me when I was in person at the hospital, and I didn’t really understand what they were on about, but the person in the hospital said to my son, do you understand it? And he went, yes, there you are, you’re sorted.”* **– Cassie**

## Theme 6 - Discussion

Participants in this study described several burdens that made trial participation feel laborious and emotionally taxing, echoing prior research on how excessive burden harms participant experience(35). Although remote tools are often promoted as convenient, flexible, and efficient, reducing participant time investment and enabling more passive data collection(7–10,70), participants frequently faced friction, constraints, and additional trial responsibilities. The cumulative effect of having “one more thing to manage” heightened emotional stress, as technical, administrative, and health-related demands became entangled. This burden risked undermining participant retention and agency, particularly when informal technology support became essential for coping. Reliance on external support networks, such as family, shifted responsibility and could create passive participant roles, while also imposing burdens on family members providing support. This dependence highlights possible design and usability flaws and raises equity concerns for participants without access to such support. The findings reveal a disconnect between the efficiency goals of remote trials and the complex, emotionally charged realities faced by participants who must simultaneously manage illness, personal life, and trial demands. While remote models aim to promote autonomy, without adequate supporting systems this autonomy can become a burden, exposing a disconnect between self-management ideals. Managing trial burdens and technology may seem minor in isolation but can become overwhelming when layered with health and day to day challenges.

## General Discussion

We conducted 14 interviews and employed thematic analysis to explore the perspectives of remote clinical trial participants regarding their involvement in DCTs. By focusing on participants’ experiences and perceptions, we gained valuable, in-depth insights into their involvement in these trials. Our findings highlight how participants typically embrace remote trials and can find meaning as partners and co-owners in trial tasks. There is also often an erosion of participant experiences, a lack of personalisation, and understanding of participants. The study sheds light on the broader implications of remote trial participation and its impact on the participant experience.

## Implications for Participants

The shift toward remote and decentralised clinical trials is reshaping the participant experience, offering clear benefits but also significant challenges. While these models improve convenience and accessibility, they can also impose social, practical, emotional, and cognitive burdens. Convenience is valued but not sufficient to sustain engagement or ensure a positive experience. Several elements central to participant satisfaction may be undermined, including compassion and personalised attention(31); relationships with staff; social interaction and trust-building(32,71); clear communication(33), regular contact and feedback(22); and a genuine sense of care(35). Participants also report practical difficulties, including issues with usability and digital literacy(22). Viewed through a behavioural science lens(57), we can see that autonomy, relatedness, and competence are all affected, core psychological needs that must be supported to maintain motivation and retention(41). If these needs are not recognised and addressed, there is a risk that remote models may feel less engaging or even alienating for some participants. To address this, DCTs must evolve to prioritise inclusive, flexible approaches and support systems that respond to diverse participant capabilities, preferences, needs, and levels of confidence, offering multiple ways to engage, support, and resolve issues efficiently. Emotional needs also matter(72) and support should extend to provide ongoing care, opportunities for connection(73), and personalisation(16). These elements can help build trust(32), reassurance(74), and a sense of connection(75,76), and contribute to positive experiences. Barriers remain, including assumptions about digital readiness(77), entrenched models, and the challenge of investing in more personalised, less scalable approaches. Yet clear enablers exist: early patient involvement through co-design(78), piloting and testing tools with diverse user groups(79) and embedding feedback loops to support continuous improvement. Addressing these issues enables a more human-centred approach that strengthens engagement and retention by reflecting what truly matters to participants. Trial designs must therefore move beyond convenience and efficiency to embed personalisation, flexibility, and holistic support throughout, laying the foundation for ongoing, tailored support aligned with participant needs.

### Implications for trial design

Given these needs, trial designs should reflect the diverse and often complex experiences of participants by embedding personalisation, flexibility, and ongoing support throughout the study. Similar to past research(15,17,18) our study highlights that DCTs address some of the challenges of traditional on-site clinical trials(80) by improving convenience for participants. However, participants differ widely in their need for reassurance, feedback, and human contact to stay engaged. Our findings show participants experience DCTs as detached and mechanistic, underscoring the need to adapt approaches for remote, technology-mediated settings. To genuinely support participants, trial design must go beyond procedural efficiency, remote clinical trial designers must also aim to prioritise personalisation and flexibility, along with coherent, ongoing support that integrates emotional, relational, and practical needs. Trial design must account for this, embedding systems that help participants feel informed, capable, and genuinely supported. Fully remote or rigid models do not suit everyone. Some participants rely on in-person interaction to build trust and feel secure, while others need regular communication to stay engaged and feel supported. Traditionally, research teams have supported retention through personalised and relational support(16,41), but decentralised models can disrupt these connections. Our research highlights the importance of viewing the participant experience holistically, recognising that clinical, psychological, emotional, and practical factors are interconnected. Participation is not just about procedural compliance; it’s also about feeling valued, safe, supported, and positively engaged throughout. To achieve this, trial design should anticipate variation across participants from the outset. Hybrid models tailored to individual circumstances(16) and consistent, meaningful interactions with study staff are key to building relationships sustaining trust and motivation(22–24). At the same time, practical constraints such as limited staff resources and operational pressures make tailored support or site visits difficult. This is where thoughtful technology design can play a crucial role, enabling flexible and responsive ways to meet participants’ needs, but the technology must be intentionally designed to support and sustain those needs throughout the trial. Here, trial design intersects with technology choices, which can help extend flexibility and responsiveness.

### Implications for the design of technology to support remote clinical trials

Technology is the primary interface through which participants experience DCTs, and can shape how they feel supported, connected, and engaged. Consistent with prior research(58,70), our findings show that current technologies prioritise data collection and efficiency (technical aspects) while neglecting emotional, relational, and humanising elements (social aspects). This creates a socio-technical imbalance, reflecting what socio-technical theory describes as a misalignment between technical systems and users’ social needs(81). In the context of DCTs, these social needs can be understood through a participant-centred lens, which emphasises involvement, communication, support, and contextual fit. Guided by socio-technical considerations, technologies should therefore integrate technical functionality with these social dimensions alongside data capture. Features such as two-way messaging, personalised communication, context-sensitive feedback, and real-time monitoring of well-being can help participants feel acknowledged and supported, aligning with broader calls for patient-centred trials(27). Emerging technologies offer further opportunities, AI can enhance communication, feedback, and relational elements with personalised communication via natural language processing (NLP)(82), and adaptive content tailored to individual learning needs(83). Virtual assistants(84), can provide immediate, tailored responses and support while escalating more complex concerns to human staff. With consent, AI can also analyse text, voice, or facial expressions to detect signs of frustration, distress, or disengagement(85), and monitor shifts in mood and health status, along with real-time measures of emotional well-being(86). Implementing these innovations may require regulatory frameworks and oversight processes to evolve to support participant-centred approaches and AI-enabled functionalities. Although AI and GenAI present significant opportunities, technology design must be grounded in behavioural science theory(87). Frameworks such as Self-Determination Theory(88) support autonomy, competence, and relatedness, which can enhance participant motivation and ultimately retention(41). Practical enablers include applying usability heuristics(89), contemporary design principles(90), and inclusive co-design with diverse stakeholders(91). Participant-facing technology in DCTs offers the chance to support diverse needs throughout the trial, acting as a true companion. Realising this potential requires technology stakeholders to rethink current approaches and intentionally design tools that sustain participant needs throughout the trial.

## Limitations and future research

The insights from our interviews provide an informed starting point rather than a final or universally applicable conclusion and serve as a foundation for further exploration. Future research should examine and test how behavioural science can guide the design of remote decentralised technologies across diverse trial contexts and populations to support participant-centred experiences. Specifically, co-design activities with diverse particpants groups and technology stakeholders need to be explored as a way to develop and refine functionality that reflects real-world use and lived experience. Further studies are required to test and adapt these findings across various trial contexts and populations. Longitudinal studies following participants throughout trial journeys can provide insight into evolving needs, while studies focused on training trial staff in remote engagement techniques can enhance relational support. Finally, integration of behavioural science frameworks into trial protocols and technology development warrants exploration to improve participant motivation and retention across diverse trial contexts.

## Conclusion

This research suggests that DCTs have long-term viability as a trial approach. They offer an effective and acceptable model, improving access and convenience while giving participants comfort, autonomy, and empowerment. However, the socio-technical opportunities and challenges in running remote DCT trials are complex. Digital exclusion can arise not just from individuals’ limited resources or abilities, but also from the design and accessibility of the technologies and infrastructures that support participation. Our findings reveal a complex and fragile participant journey shaped by the remote setting, technology, and trial participation. While remote trial models reduce some burdens, they can also introduce psychological, emotional, and practical challenges. In this context, strategies that have proven effective in traditional site-based trials, including approaches to supporting and engaging particpants, may not translate effectively to decentralised trials. These challenges risk undermining the overall experience if left unaddressed, especially in the context of isolation and ill health. Clinical trials face the challenge of balancing the collection of high-quality data, ensuring regulatory compliance, maintaining participant safety, managing participant burden, and upholding scientific integrity. Without intentional focus on participants’ needs, a participant-centred experience can be overlooked. Convenience alone does not guarantee a positive experience. Emotional, psychological, and practical needs shape how participants interact with trials. Designing with these factors in mind is essential to building models that are both effective and sustainable. Given these complexities, a human-centred approach that emphasises participants’ experiences is essential in decentralised trials. Approaching technology as a relational partner and trial companion, while integrating particpants insights into its design, can strengthen engagement, trust, and retention. This study highlights the lived experiences of trial participants. It calls on stakeholders to respond to the findings, integrate them into design, and consider how technology might best support participant-centred experiences.

## Acknowledgements

We would like to thank all the participants interviewed for their invaluable contributions to this study and extend our gratitude to all individuals who assisted in participant recruitment for the study.

## Author Contributions

EG, and CL conceived the study. EG, CL, and CH contributed to protocol development and obtained ethics approval. EG researched literature, recruited participants, and conducted data analysis with review from CL and CH. EG wrote the first and subsequent drafts of the manuscript. All authors reviewed and edited the manuscript and approved the final version.

## Ethical approval

The ethics committee of University College Cork approved this study (SREC Log umber: 2024-021)

## Consent to participate

All participants provided written informed consent prior to engaging in this research.

## Data Availability

In line with university guidelines and the participants’ signed consent, we ensure that all data handling is conducted in accordance with ethical guidelines and confidentiality, with the study participants not granting consent to make the dataset available.

## Abbreviations

DCT: Decentralised Clinical Trials

## Multimedia Appendix of supplementary files

Supplementary Material 1: Interview Guide

Supplementary Material 2: Example Thematic Data Analysis Supplementary Material 3: Online Screener

Supplementary Material 4: Findings – Themes and Sub-themes

